# Gaining Control of Combination Cancer Treatment Risk by Incorporating Cost and Value Data into the Drug Selection Process *at the Point–of–Care*

**DOI:** 10.1101/2022.02.13.22270914

**Authors:** Richard L. Nicholas

## Abstract

The use of combination therapies*, as well as FDA-approved drugs for off-label indications, to treat advanced cancer, is widespread. While much is known about their clinical effectiveness, there exists no examination of the relative cost of novel multidrug combinations vs. traditional available therapy options, or study as to how knowledge about comparative therapy costs at the point-of-care can be leveraged by doctors, health systems, and payers. We found that:

combination multidrug cancer regimens may be *less costly* than monotherapies or other standard options;
novel, multidrug combinations are often *better financial values* than monotherapies or other standard options;
having treatment cost and value data, at the point of care, enables the prompt selection of more cost-effective medications and the avoidance of expensive low-value therapies that are financially wasteful.

We conclude that the effectiveness of value-based purchasing initiatives may be amplified if physicians and payers use comparative treatment cost/value data to enhance their cancer drug-selection decision making.

* Including combinations of immunotherapies, chemotherapies, targeted drugs with distinct mechanisms of action, etc.

**Study Highlights:** What Is The Current Knowledge On The Topic?

☑ *The effectiveness of molecularly targeted multidrug therapies used to treat advanced cancer is well established;* ^*1-4*^ *that few clinicians are aware of the cost of the medications they prescribe, or which are more cost-effective, deliver a better return-on-investment or represent a financial value;* ^*8*^ *and, that it is intuitive to believe that a combination of multiple high-cost medications is more expensive than a single-drug or other standard therapy options*.

What Question Did This Study Address?

☑ *Although studies on the clinical impact of multidrug cancer treatments abound*, ^*1-4*^ *there are no examinations of the relative cost or value of combination therapies vs. that of traditional monotherapies, or how knowledge of how this data can be used in practice. A systematic method to calculate, evaluate and compare the relative cost of mono-therapies, 2- and 3-drug combination cancer therapy options is presented for use by physicians, health systems and payers to better manage their oncology specialty pharmacy spend and drive better medical outcomes*. ^*3*^

What Does This Study Add To Our Knowledge?

☑ *We show that multidrug cancer therapies are not necessarily more costly than single-drug or other standard therapy options; and that furnishing physicians and payers with comparative treatment cost and value data to augment their complex medication selection decision making enables them to identify drugs that are a value, avoid those that are wasteful, and create better targeted novel combination cancer therapies that represent a value, which incorporates both clinical and financial aspects*.

How Might This Change Combination Therapy Drug Selection Or Value-Based Oncology Management?

☑ *Clinicians have the tools, information, and data with which to confidently prescribe novel drug combinations that customize molecular targeting, and lower treatment costs. Payers now have a framework within which to drive value-based purchasing to gain control of their oncology specialty drug risk. Patients will benefit from more personalized, efficient and effective therapies and less financial toxicity (i*.*e*., *distress)*.

## Introduction

The use of customized combination drug therapies having a molecular rationale is now common among leading oncologists. While they have been proven to be effective, little is known about their comparative cost or value. This study provides a methodology by which to measure and compare the cost and value of mono- and multidrug cancer treatment options; and demonstrates how having this data at the point-of-care can impact patient, physician and payer decision making to better identify therapies that are a value, and to avoid those that are not.

## Method

This study presupposes that the science supporting the clinical effectiveness of customized multidrug cancer therapies is proven. ^1-4^ and that the associated measurement tool is accurate in determining the degree to which a cancer therapy option addresses a patient’s molecular cancer profile. ^5-7^ In choosing a study methodology, we considered the validity, appropriateness, infirmities and implications of many healthcare economic analysis instruments. Given our narrow scope, and the power of the underlying clinical measure as an effectiveness predicter, it was not necessary to consider a wider scope of treatment costs, long-term medical expenses, or other health outcome measures to fairly illustrate the comparative financial impact of one treatment vs. another. As such, we coupled a cost indicator with the clinical effectiveness report results to create a *cost/value* index that can be used to identify best treatment values (outcomes/cost) and avoid wasteful low-clinical-value drugs.

### Study Data

This study’s clinical effectiveness data was obtained from *CureMatch, Inc*. in the form of reports generated by its AI-powered therapeutic decision support platform. This tool identifies drugs known to act against specific cancer mutations then scores and ranks drug combinations that best target a tumor’s actionable variants (i.e., the score correlates to progression-free and overall survival rate). ^5-7^ This study can be replicated using data from the same source or using combination drug/mutation-matching data from another source.

The 10 real-world case studies featured in this analysis were selected by employing criteria intended to identify representative advanced cancer examples that would likely warrant a clinician’s consideration of a high-cost combination drug treatment (e.g., stage 3 or 4, multiple actionable biomarkers, etc.). Case studies were selected before either the drug pricing or value indexing methodologies were developed, and before the cost of any drug treatment option could be calculated. An N =10 sample size was determined to be more than sufficient to produce statistically significant results for our illustrative purposes.

### Combination Treatment Cost Calculation

To arrive at the 3-month cost of the various combination cancer treatments we followed these steps:

1. *Identify each of the unique drugs that was used in a combination treatment and determine if a generic form of the drug was readily available at the time the case study that it was used in was undertaken*. This study involved 36 unique drugs, 32 (89%) of which were patent-protected brands at the time of the case study. Only 3 drugs, each chemotherapies, had a generic form; 1 is a brand-transitioning-to-generic.
2. *Establish a dose adjustment protocol for 2- and 3-drug combination treatments*. Each cancer drug dose may be substantially reduced when drugs are used as a part of a multidrug therapy. We applied a reduction of 50% for 2-drug combinations and a reduction of 67% for 3-drug combinations. For some combinations dose reductions may be more or less.
3. *Estimate the most probable adjusted dosage for each combination treatment type (i*.*e*., *1-, 2- or 3-drugs) and evaluate each drug to determine if it is possible in practice to fashion the required adjusted-dose regimen*. To endure that these dose adjustments can be made in real-world clinical settings, we evaluated the strength, size, packaging, half-life, dose levels and cost of each drug, and factored the common practices used to expand dosing variety (e.g., frequency, interval) into the calculation (see *Appendix A: Combination Drug Dosing Challenges*). The variety of each drug’s dosing options was classified as high, moderate or low; where moderate denotes more than ample variety. We classified 21 (58%) of the 36 drugs as high variety; 14 (39%) as moderate variety and 1 (3%) as low variety; evidencing abundant (97%) moderate- to high-level variety.
4. *Select a brand and generic drug pricing basis and establish and apply suitable discount levels*. Drug prices are impacted by their many suppliers, buyers, and sellers. Many use different price schemes. Unlike most products, drug prices are complex and illogical (e.g., they don’t always go up each time they change hands) and rarely reflect what is actually paid for them (see *Appendix B. Pharmaceutical Pricing)*. We used the pricing benchmark employed by the pharmacy benefit managers (PBMs) that administer pharmacy plans for payers: the average wholesale price (AWP). We priced each drug to reflect the full monotherapy dose for the indication that the drug is most commonly used, based on pricing that reflects the most cost-efficient package option (when unit price is volume sensitive). A drug’s AWP is discounted based on a payer’s size, drug spend, formulary placement, etc. We applied a 15% discount to brand drugs and an 80% discount to generics; levels typical in the payer market. The results appear in Table 1 below.

**Table 1:**
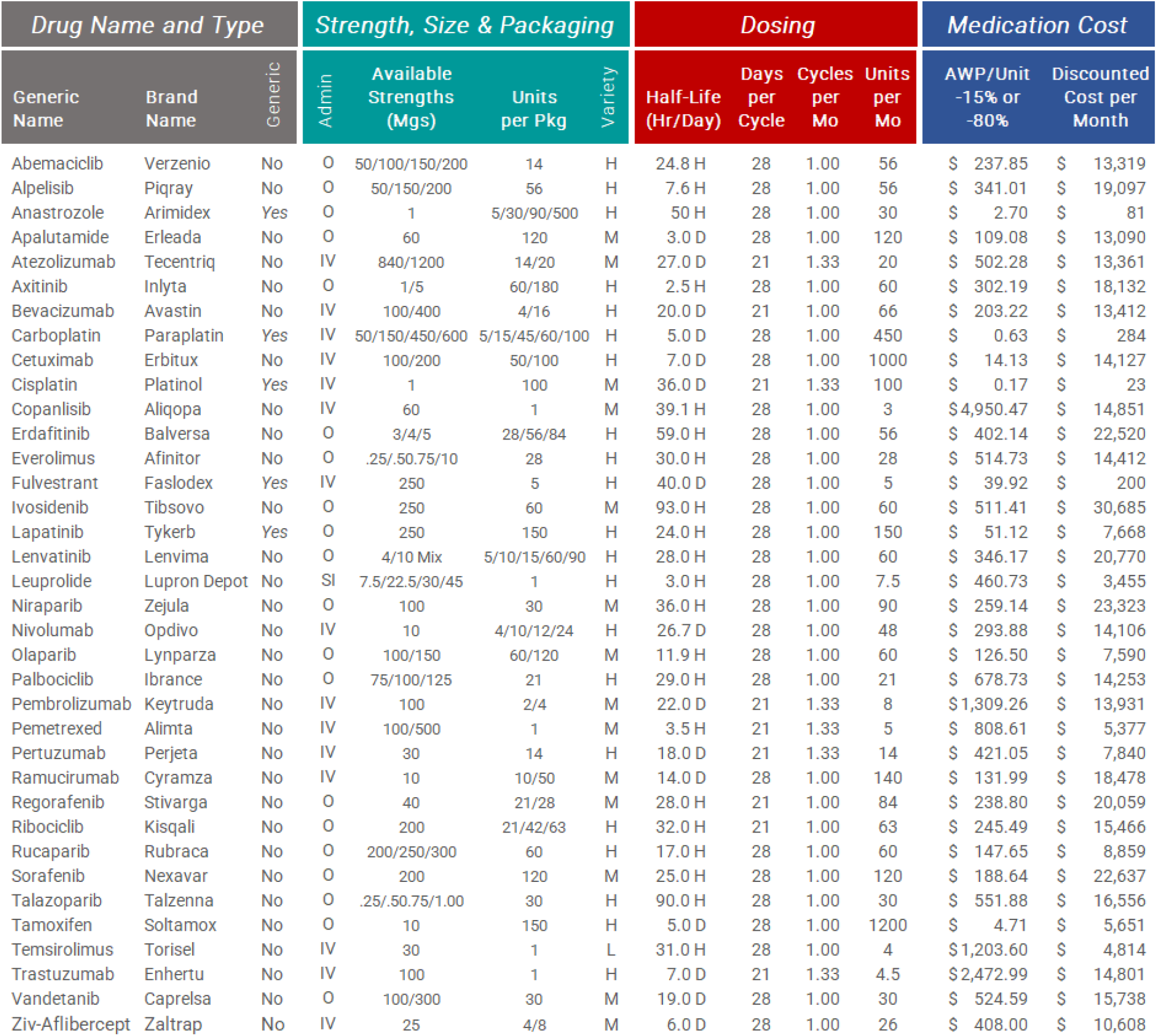
Treatment Medication Costs/Dosage Adjustment Variety
5. *Calculate the cost of using each drug for 1 month, at full strength; and then the cost of each 2- and 3-drug treatment option, applying appropriate dosage adjustments*.

The price for a drug’s full monotherapy dose for 1 month was used as the basis upon which to calculate the 3-month drug cost of the 90 scored and ranked combination drug treatments. To be cautious, we increased the drug cost basis by 20% (from 33% to 40% and from 50% to 60%) for 2- and 3-drug treatments respectively, to compensate for any inefficiency owing to dose size, packaging or strength.

### Report Elements, Format and Interpretation

The following is intended to add clarity and context to the case study clinical effectiveness exhibits below:

- There are approximately 300 FDA-approved drugs that are used in the US for the treatment of cancer. When employed concurrently, there are ∼4.5 million possible 1-, 2- and 3-drug combinations of these medications.
- A *Pathogenic Marker* is a genetic alteration known to cause and/or be associated with cancer whereas an
- *Actionable Marker* is a genetic alteration that has a known response to an existing FDA-approved drug.
- *Combinations Considered* is the number of 1, 2, or 3 drug combinations that were subject to analysis.
- *Relevant Combinations* is the total number of drugs that were found to be relevant to the patient based on their tumor’s unique molecular profile, and that were considered in the analysis.
- *Treatment Score* reflects the degree to which a given therapy option addresses a patient’s molecular cancer profile (a higher score correlates with a better outcome). ^6-7^ Drugs are selected for their potential to target mutations. The score may be used to compare regimens within or between profiles and cases.

The standard report features only the 3 highest-scoring 1-, 2- and 3-drug treatment options. Below, *treatment cost/financial value* results (right, in blue) are added to *comparative clinical treatment* results (left, in gray).

- The *Full Dosage Cost* is the sum of the full cost of all the drugs used, at full dosage, for 1 month.
- The *Adjusted Tx Cost* is the 3-month cost of treatment, after appropriate dose adjustments.
- *Best value* is defined as the lowest therapy cost relative to the Rx combination/mutation-matching score and the *Value: $/CM Point* is largely a comparative cost/value metric of predicted targeting effectiveness.

The formula is: Value Index ($/CM Point) = (3-month-adjusted dose) divided by the CureMatch score (%). In the example, the full cost of one month of cisplatin plus palbociclib plus bevacizumab is $27,689. After adjusting the dose and the 3-month interval, the cost is $33,227 for 3 months of therapy. As the matching score is 83% for this example, the cost per CureMatch score percentage point (Value Index ($/CM)) is 400, which suggests a better value than the equally-scoring next combination having a Value Index of 404 $/CM. The highlighted option is also of better value than the lower scoring 2-drug combinations, such as cisplatin and sorafenib (Value Index = 769)

From a clinical perspective, the integrated case results above (Figure 1) should be interpreted as follows:

- NGS testing identified 3 actionable pathogenic markers;
- 23 drugs were molecularly matched to an actionable marker (all but 3 would be used off-label);
- 2,024 relevant 1-, 2- and 3-drug treatments were identified, evaluated, scored and ranked; and
- each of the top 3-drug treatment options look very promising with scores ranging from 78% to 83%.

**Figure 1:**
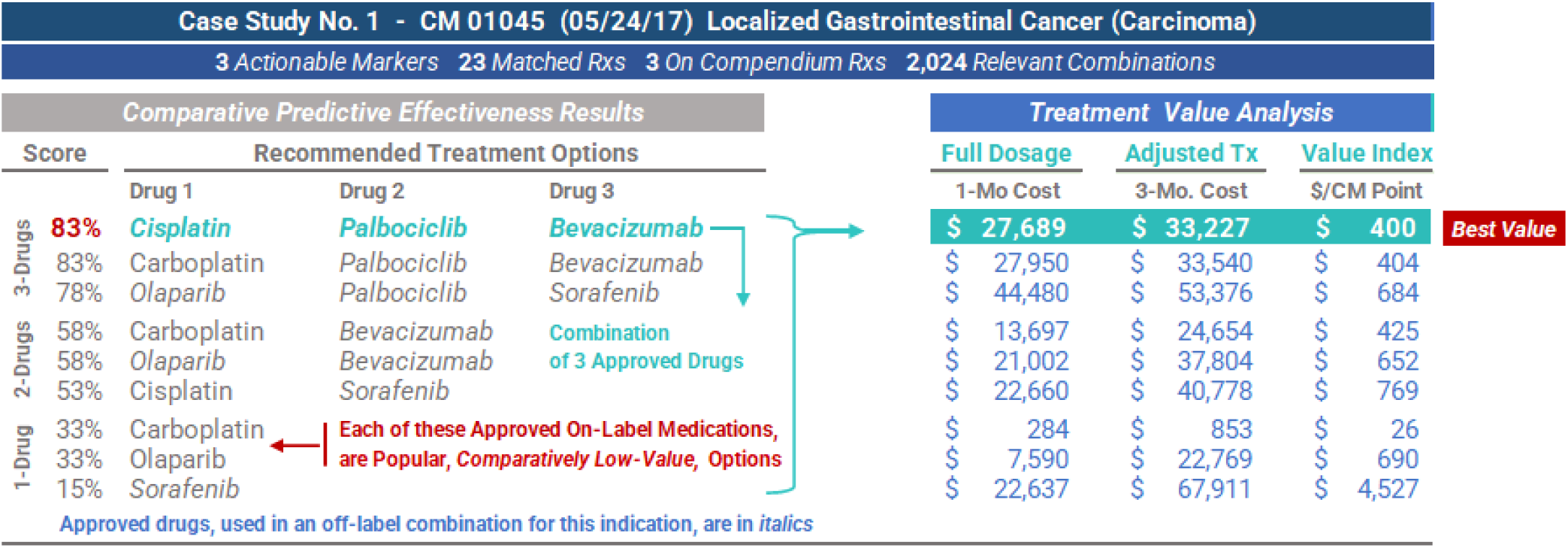
Comparative Clinical Efectiveness/Value Analysis - Case Study No. 1

From a financial perspective, the integrated case results above should be interpreted as follows:

- the 2 top-ranked combination options are essentially the same (from a financial perspective);
- with a regimen cost of ≈ $34,000 and a cost/value rating of $410 ($34,056 ÷ 83), they are *best values;*
- each monotherapy is a standard option for this indication, and, in this example, each is a *lower value* option.

Interpreting the results of the case study above is instructive as it validates the structure and methodology of our study. Having comparative treatment cost/value data at the point-of-care facilitated the identification of high-cost, low-clinical-value therapies, and enabled the identification of the top 3-drug combination options as *best values*. As these findings are counter-intuitive and otherwise unapparent but for this type of analysis (e.g., many high-cost drugs should be *more costly* than a single drug), they evidence that our methodology is correct.

## Results

The following study findings are presented in a logical sequence, without bias or interpretation, and intended to help the non-clinically trained executive reader better understand each finding, in its proper context.

Counterintuitively, our findings show that when multiple cancer drugs are used concurrently, with the dosage of each adjusted correctly, combination drug therapy is often *less costly* than many single-drug treatments. We note that 35 (97%) of the 36 drugs used offered more than adequate variety to facilitate the dose adjustment.

We found that 7 of the 10 *best value* treatments were 3-drug combinations of singularly high-cost drugs; the others being 2-drug options. Multidrug therapies were often found to be *better values* than single-drug ones.

Our findings showed many expensive, yet very popular, single-drug therapies to be of *low-clinical-value* (re: the subject indication). The purchase of these drugs would therefore be wasteful and should be avoided.

Of the study’s 90 combination therapies, 66 (73%) used at least 1 drug off-label; creating a novel combination.

We illustrate how equipping physicians and payers with comparative cost/value treatment data at the point-of-care may be essential to more accurately identifying therapies that avoid waste, improve care, and drive value.

A representative sampling of some of the study’s findings, at the case study level, follows:

### Case Study 3

The 6 top-ranked treatments scored 82 – 84%; the *best value* being the <u>least costly</u> of these. Note the enormous difference in the cost between the 2 low-clinical -value (17%) options. (Figure 2)

**Figure 2:**
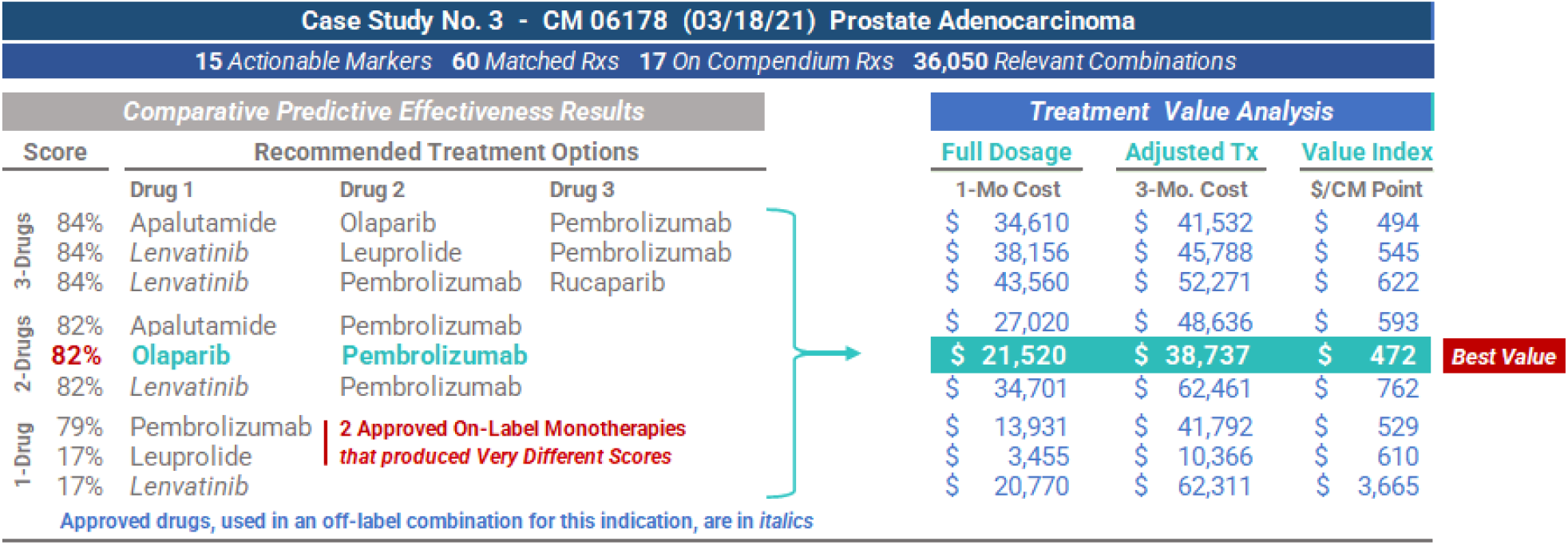
Comparative Clinical Efectiveness/Value Analysis - Case Study No. 3

### *Case Study* 4

A 2-drug combination represents the *best value*, with a comparatively high score of 74%, creating a significant financial savings opportunity vs. the top-scoring 3-drug options. (Figure 3)

**Figure 3:**
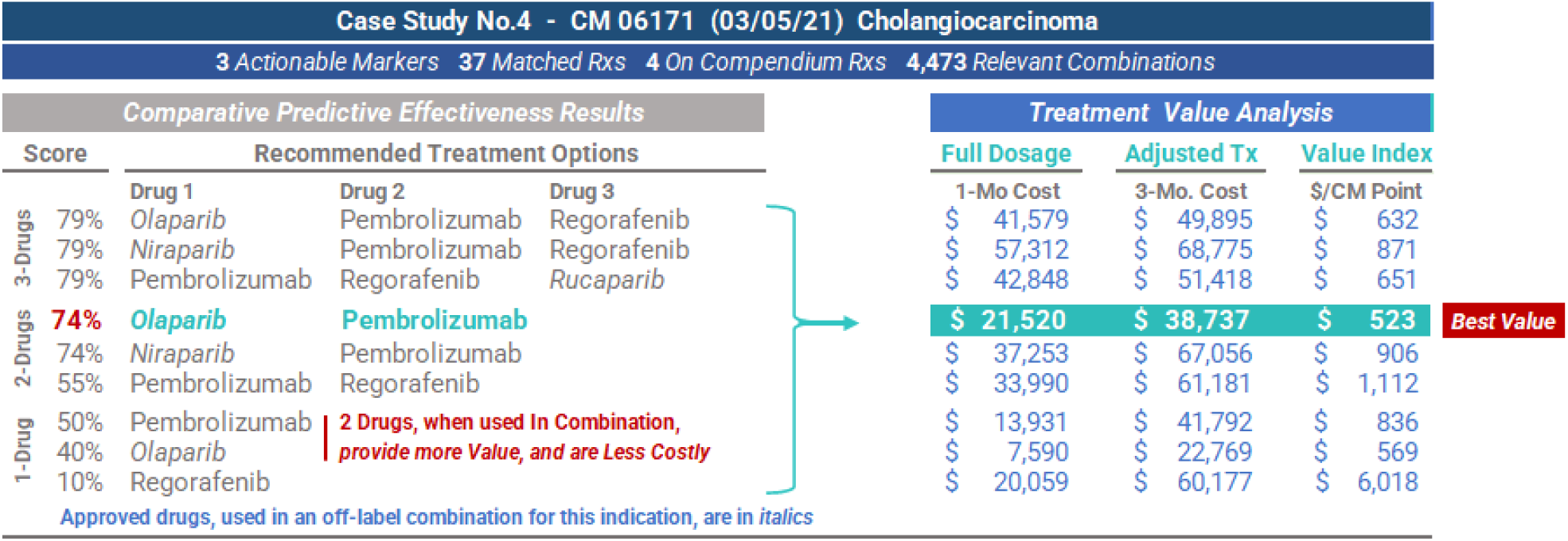
Comparative Clinical Efectiveness/Value Analysis - Case Study No. 4

### Case Study 8

The best scoring option, a 3-drug off-label combination, represents the *best value*, achieving a comparatively high score of 62%, while creating a significant financial savings opportunity vs. all of the scored options. Note that each of the 2 low-clinical-value options are far more costly than the best value (with 1/5^th^ the score). (Figure 4)

**Figure 4:**
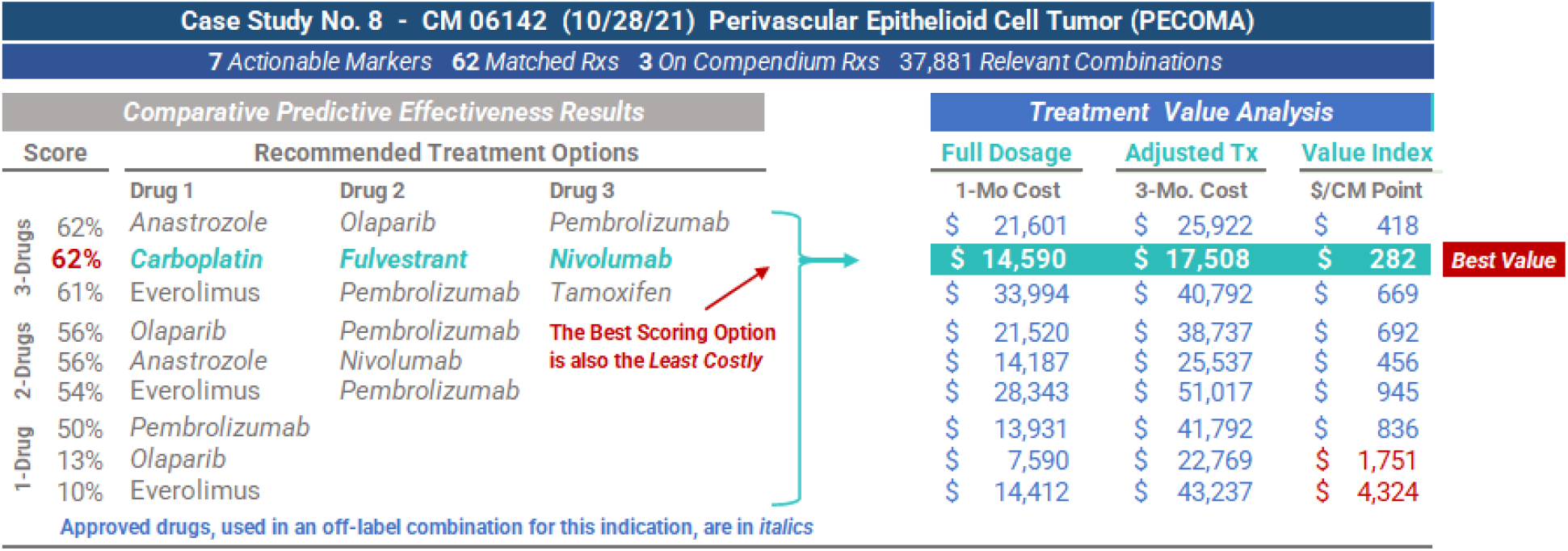
Comparative Clinical Efectiveness/Value Analysis - Case Study No. 8

In summary, these case study findings (counterintuitively) show that 2- and 3-drug combination therapies are often times less expensive, and represent better financial values, than many popular single-drug options (several of which were found to be of low-clinical-value as it relates to the subject indication). These results show that having point-of-care cost/value data can help to identify treatments that avoid waste and those that are values.

## Discussion

The use of customized combination cancer therapies is now commonplace and, while often very effective, little is known about their comparative cost and value. Precision medicine treatment involves hundreds of drugs and requires the evaluation of thousands of possible combinations; a task that exceeds human cognitive capability without AI assistance. Distinguishing between drug combinations that are toxic or of low-clinical-value, and those that are likely to work and be cost-effective, is also difficult. These challenges can be overcome, however.

Few clinicians are aware of the cost of the drugs they prescribe ^*8*^ ; while others have misconceptions about them. Both often result in high-cost, ineffective treatment. This study evidences that combination cancer therapies are often *less costly* and a *better value* than alternative monotherapies; something that is counterintuitive and unlikely to be identified without an AI-supported tool, and treatment cost/value data. Clinicians *alone* cannot be expected to create complex drug therapies that are both clinically effective *and* a value. We demonstrate that when equipped with comparative treatment cost/value data, clinicians and payers can identify multidrug cancer therapies that are better, less costly and a greater value vs. popular, yet low-clinical-value, monotherapies and other standard therapies.

We provide a methodology by which to measure and compare the cost and value of mono- and multidrug cancer treatment options. Physicians and payers can employ this comparative treatment cost/value index to augment and enhance their drug-selection decision making and amplify the effectiveness of their value-based purchasing.

## Conclusions

The employment of advanced DNA sequencing data and AI-supported systems to create customized molecular tumor matching treatment options is becoming the standard of cancer care. ^5-7^ As the widespread use of these technologies increases, a genuine opportunity arises to replace *waste* with *value* re: combination cancer therapy.

Together, physicians and payers can use this new technology to guide care, reduce toxicity, improve outcomes, control utilization, avoid waste and better manage their finite resources by expanding their organization’s value-purchasing to include expensive specialty cancer drugs. Uninformed, high-consequence oncology prescribing is not consistent with value-based care. This study demonstrates that there is no longer a reason for it to continue.

## Key Terms and Concepts

This section is intended to provide the reader with an understanding of, and context for, some of the important terms, concepts and constructs discussed herein that may not be common knowledge to non-clinical experts.

### Targeted Cancer Therapy

*Targeted cancer therapies*, aka *molecularly targeted drugs*, are deliberately chosen precision medicines and agents that interfere with specific molecular targets that are involved in the growth, progression and spread of cancer. They do this by leveraging data and information about a tumor’s unique genomic profile, in contrast with chemotherapy that acts rather indiscriminately on all rapidly-dividing normal and cancerous cells.

### Combination Drug Therapy

*Combination drug therapy* is a treatment modality that combines 2 or more therapeutic agents to enhance efficacy (vs. a single-drug approach) by targeting key pathways in a synergistic or additive way or by providing multiple opportunities for a response to an agent. First employed in 1965 to successfully reduce tumor burden and prolong remission in leukemia patients, combination therapies have been used to treat many diseases, most notably HIV/AIDS, since 1996 when the FDA-approved the drug cocktail became the new standard of care for HIV. FDA-approved multidrug therapies to treat cancer date back to the 1990’s. The safety, efficacy and cost-effectiveness of combination therapy is well documented ^1-4^ and, as such, they have become a cornerstone of cancer treatment.

Multidrug cancer treatments have proven to be so successful that drug makers are researching, and marketing, pre-packaged combination therapies. An example is OPDIVO^®^ + YERVOY^®^, an FDA-approved 2-drug immunotherapy combination used to treat some mesotheliomas. Each drug helps active T cells provide protection from foreign threats in different yet complementary ways. OPDIVO^®^ (*nivolumab*) helps active T cells identify cancer cells that are hiding; YERVOY^®^ (*ipilimumab*) helps generate more active T cells to seek/destroy more cancer cells.

### Drug Repositioning / Off-Label Use

*Drug repositioning*, also known as off-label drug use, is a therapeutic approach whereby current pharmaceutical agents primarily used to treat one disease are used to treat a different disease. A good example of drug repositioning is, Rapalog, an immunosuppressant that is primarily used after an organ transplant to prevent graft rejection, which has shown to have cancer prevention properties, owing to its mTOR inhibitory ability.

Apart from pre-packaged FDA-approved combination therapies that address specific gene mutations (e.g., those targeted by OPDIVO^®^ + YERVOY^®^), off-label drug use is associated with the use of FDA-approved drugs in customized combinations intended to address gene mutations in novel ways; the premise being that there is a greater likelihood that a novel combination therapy will yield a better outcome the first time vs. repeated attempts using standard treatments (often at a lower overall long-term cost, targeting one marker at a time, on average). While approved drug safety protocols and known pharmacokinetic profiles are evaluated and expertly addressed, it is not feasible to have such clinical trial results for millions of combinations possible from the ∼300 anti-cancer drugs. Hence, some oncologists are reluctant adopters who reserve this practice only for late-stage patients, when standard-of-care monotherapies and pre-packaged on-label combinations have failed or when genomic markers indicate they may be effective when the molecular match is exact and clinically documented as such.

### Effectiveness

The terms *effective* and *ineffective* are used herein to refer to the degree to which there exists a molecular rationale to prescribe the treatment; and not intended to describe the effectiveness of the therapy itself.

### Novel

The term *novel* is used herein to refer specifically to a *combination* of FDA-approved drugs, any of which may be on- or off-label for the given disease. It is not intended to infer that each drug by itself is novel.

### Value and Value-Based

The term *value-based* is used to describe healthcare payment, reimbursement and delivery frameworks aimed at improving quality and bettering outcomes for patients; with physician payment and incentive models that reward quality improvement and health outcomes over volume; appreciating the reality that the financial resources of all payers are finite. Therefore, *value* can be defined as health outcomes per unit of cost. In the context of this study, value is also indicated by an index: *tumor matching score per unit of (3-month) drug cost*.

## Limitations

Healthcare economic analysis often examines the *full financial impact* of one drug regimen vs. another, typically considering a wide scope of therapy expenses other than just drug costs (e.g., NGS testing, drug administration, professional services), health outcomes data, QALY measures, predicted long-term medical expenses, etc. The narrow purpose of this analysis is to show that intuition-based treatment combination drug selection logic (e.g., multidrug therapies must be more costly than monotherapies), and the limits of human cognitive ability, both warrant the augmentation of high-stakes, complex clinical decision making with not only AI-enabled clinical support but also point-of-care treatment cost/value data. In this limited context, there is no need to examine the full financial impact of each therapy option, but only to fairly illustrate the *relative value difference* between them.

With respect to the calculation of the cost of each therapy option, some believe that using the drug price in effect at the time the drug was dispensed (the case study date) would be more precise than using today’s price. We did not have access to global historic drug prices, however, we found no evidence to indicate that the relative price differential between therapy options changed much, or at all, to materially impact any of our findings.

There are cases where not all drugs in a combination can be adjusted equally (e.g., some may be escalated to higher dosage levels over time while others may be reduced further, based on the drug, patient clinical presentation, etc.)

As the study is US-centric it may be less applicable in nations with different health system funding arrangements.

While these results are meaningful, more research is needed to fully examine the extent to which providing comparative point-of-care drug cost/value information can reduce treatment waste and drive value purchasing.

## Data Availability

All data produced in the present study are available upon reasonable request to the authors

## Data Availability

This analysis was prepared by the *TPA NETWORK Research Consortium*. For more information or data about this study or topic, please contact the author, Richard L. Nicholas, at or (858) 395 – 4114.

## Disclosures

The *TPA NETWORK Research Consortium*, the primary study funder, serves as a healthcare industry advisor and payer product development consultant to *CureMatch, Inc*. which provided indirect funding for a limited portion of this research. No funding source had any role in any aspect of this analysis and the existence of any relationship does not constitute a conflict of interest, or otherwise bias the impartiality, or compromise the integrity, of this study.

Below are several studies that provide clinical support for the science and technology discussed herein, including the landmark NIH-funded I-PREDICT [study](https://pubmed.ncbi.nlm.nih.gov/31011206/) recognized by the *American Society for Clinical Oncology* as a major achievement in cancer research in 2017 and featured in Fmr. NIH Director Dr. Francis Collins’ 04/19 [blog](https://directorsblog.nih.gov/2019/04/30/personalized-combination-therapies-yield-better-cancer-outcomes/#comments).

## Appendix A: Combination Drug Dosing Challenges

As a point of distinction, *dose* refers to a specified amount of medication taken at one time and *dosage* and *dosing* refer to the specific amount, number and frequency of doses taken over a specified period of time.

The dose of a drug modulates whether patients will experience optimal effectiveness, toxicity, or no effect at all. With respect cancer drug dosing, the goal is to maximize efficacy and minimize toxicity. Traditionally, dosages have been calculated based largely upon a patient’s specific situation e.g., body surface area/mass (a factor of weight/height), age, gender, smoking, liver and kidney function, disease-specific considerations, possible drug/food interactions, etc. This approach is based on the theory that larger patients have a greater volume of distribution and higher metabolizing capacity, thereby requiring more of a drug to achieve the same effect.

Traditional dosing has been *normalized* to *minimize* interindividual variation. Correct dosing of cancer drugs is more complicated however as side effects are a key reason for patient nonadherence / early discontinuation. Moreover, dosing of *combination* cancer therapies can be challenging as it requires dose reductions to spare normal cells, while simultaneously creating the desired cytotoxic effect on the targeted cancer cells. This is especially true when two or more drugs of the same class are used, or when there are overlapping targets.

Informed, forward-thinking oncology practices are adopting personalizing (adjusted) dosing based on individual pharmacokinetics (i.e., the movement of drugs within the body); a practice that is consistent with the underlying premise of personalized/precision medicine which is to tailor treatment and care to the individual patient and their specific disease. *It is not enough to select the right drug for a patient; it is critical that the dosage be correct*.

In the treatment of cancer, combination drug therapy involves lowering the dose of 1 or more component drugs. While not universal, it is a common practice in 2-drug combinations to start with ∼ 50% of the usual dose of each drug and for 3-drug combinations at ∼ 33% of the usual dose noting that lower dosing often does not fare worse than higher dosing or alter efficacy. ^9^ Many cancer drugs are formulated, sized, packaged and priced to enable a wide variety of precision dosing options and make it easier to develop an optimal personalized dosing regimen.

Beyond clinical considerations, given the exceedingly high cost of many cancer therapies (e.g., $30,000+ /mo.), drugs that are inappropriately dosed create significant, avoidable medical expense. Waste associated with non-optimized drug therapy in the US is believed to be a half trillion dollars per year; ≈ 15% of our healthcare budget. Drug regimens must be more precisely tailored to each individual. Cancer — where nonadherence and early medication discontinuation pose real, significant, and unnecessary financial burden — is no exception.

Counterintuitively, this study evidences that the use of expensive cancer drugs does not need to result in an excessively high treatment cost. even in combination, *when the drugs are selected and dosed correctly*. These high-cost medications often work to lower the total cost of care; thereby creating a genuine value. To accomplish this, combination drug treatments must be effective for every patient; i.e., they must be accurately dosed.

Many pharmaceutical manufacturers have recognized that certain of their products were sized, packaged and/or priced inefficiently (at least for the US market), such that it made precision dosing a challenge in practice. Most now offer a variety of dosing options; in part, to make their products attractive for combination and off-label use. There are several practices and techniques that may be employed to more precisely tailor a cancer patient’s dosing regimen to support novel multidrug combination treatment and make these therapies able to be of benefit to a wider patient population. These include pill and package splitting, dose cycle alteration, the use of loading doses, fractionated dosing, dose rounding, split fill dispensing, treatment sequencing and scheduling adjustments, etc. Each is discussed in detail below in *Appendix B: Pharmaceutical Pricing*.

### Therapeutic Index and Combination Drug Treatment

*Therapeutic index* refers to the measure of the degree of fluctuation a drug is known to have over a dosing interval before it results in toxicity due to high peak concentrations. It is the ratio between the toxic dose and the dose at which the drug becomes effective. A drug having a *low* therapeutic index requires concentrations to be maintained within a *narrow* therapeutic range. This narrow therapeutic window typically limits the dose that can be achieved as a slight dosage variation may induce an adverse drug reaction or treatment failure. A drug having a *high* or *wide* therapeutic index is preferable to one having a low one as a patient would have to take a much higher dose of such a drug to reach the toxic threshold than that required to elicit the therapeutic effect. Most chemotherapies have a narrow therapeutic index. Unlike immunotherapies that require continuous dosing however, some cancer medications are dosed cyclically and can be adjusted based on clinical pharmacokinetic analysis performed on multiple blood samples drawn over a period of time. These wider therapeutic index drugs can often be given at intervals that exceed the drug’s half-life without increasing toxicity.

By reducing the dose of the drugs used in a combination therapy — and leveraging the precise individualized dosing enabled by the specificity with which certain therapies deliver potent targeted cancer agents — the *composite therapeutic index* of the combination treatment may possibly be widened, thereby enhancing the therapy’s utility, and expanding its ability to provide clinical benefit to a much broader patient population.

### Evolving Drug Dosing Constructs

In an ideal world, drugs would be available individually, in many different dose sizes and strengths and priced accordingly. As this is not the case, for clinicians and oncology pharmacists to achieve a desired reduced dose, challenges may need to be overcome. Here is how these challenges may be effectively addressed.

#### Package Sizing and Pack-Splitting

While it is true that certain drug packaging can simultaneously drive-up costs and deter precision dosing, many medications are packaged to facilitate reduced dosing over time. By example, by combining different dose sizes in the same blister pack, Lenvima^®^ (*Lenvatinib)* was originally packaged in 4- and 10-mg capsules to achieve a 24 mg dose to treat thyroid cancer (with similar packaging for 20-, 14- and 10-mg daily dosages). After being approved for other indications, 18-, 12-, 8- and 4-mg dose options were created. As many blister packs contain doses of different strengths, *pack-splitting* can facilitate precise dosing and reduce waste. Beyond its clinical value, multiple dosing options create an opportunity for substantial payer/patient savings.

#### Drug Half-Life and Dose Cycle Alternation

The half-life of a drug is an estimate of the time it takes for the drug’s active substance in a patient’s body to be reduced by half. Drugs having a shorter half-life tend to act quickly, although their effects may wear off rapidly, often resulting in the need to be taken several times a day. Drugs having a longer half-life may take longer to start working, although their effects may persist longer, resulting in a need to be dosed once a day, week, month or less frequently. A drug’s half-life varies based on many drug- and patient-specific variables.

Given the half-life of many cancer agents, small differences in daily doses generally don’t produce clinically meaningful results in the context of steady-state daily dosing. As such, minor drug regimen changes to reduce the dosage and mitigate waste are viable. Typically, this is done by taking one larger pill/capsule a day vs. 2 smaller ones; adjusting daily doses over a few days (e.g., 1 dose for 5 days and another for 3 days); extending dosing cycle intervals, etc. Most state laws accommodate practices to enable for patient-specific drug regimen tailoring. It also presents a real opportunity to achieve substantial payer and patient savings.

#### Loading Dose

*Loading dose* refers to an initial high dose of a drug that has a long half-life that is given at the beginning of a course of a treatment to allow for dosing once, twice or three times daily as the large degree of fluctuation over the dosing interval does not result in toxicity due to high peak concentrations. When done, only a low maintenance dose is required to keep the amount of the drug in the body at the appropriate therapeutic level (otherwise, it would take longer to reach the appropriate amount of the drug in the body). Drugs that lend themselves to this practice have a wide therapeutic index, which enables dosing at intervals longer than the drug’s half-life. In practice, this can result in more precise dosing, less medication waste and reduced cost.

#### Fractionated Dosing

Many cancer drugs (having a wider therapeutic index) are dosed cyclically, making them candidates for adjustment based on regular clinical pharmacokinetic analysis, customizing the treatment by changing the dosing schedule. *Fractionated dosing* is the process of dividing one dose into multiple fractions to change dose intensity while reducing peak concentration, (to reduce toxicity and prolong exposure to the drug to ensure that a greater number of cancer cells are impacted). It has been shown to improve tolerability without impacting antitumor activity, thereby potentially widening the therapeutic window of many cancer agents.

#### Pill Splitting

*Pill splitting* is the practice of cutting a pill in half to adjust a dose and/or to reduce costs (by up to 50%) by purchasing higher-dose pills. It is a common insurer- and FDA-approved practice. Film-coated pills, capsules and extended-release tablets cannot be split; and many dosage forms cannot be crushed or compounded. While tablets that do not include pill-splitting information in the label have not been evaluated by the FDA to ensure that the resulting halves contain the same drug content or will work the same way as the entire pill, it is a misconception that only scored pills may be split. Depending upon the desired objective, exact equal doses may not be of clinical significance: splitting pills *with a long half-life and wide therapeutic index* typically poses little risk. Pill splitting should be doctor approved and is best done with a common pill-splitting device.

#### Split-Fill Dispensing

Oral chemotherapies are often perceived to be safer and/or less toxic than their intravenously delivered counterparts; a perception that is inaccurate. In fact, many patients stop taking oral medications long before the initial monthly treatment regimen is completed, or drug supply finished. Whether this is the result of intolerable side effects or unrealized therapeutic benefit matters little, considerable medication waste and enormous financial consequences ensue, especially when high-cost specialty cancer drugs are involved. *Split-fill dispensing* is when high-cost drugs having pronounced side effects can be tried for a short period to confirm tolerance and effectiveness before a full supply is dispensed. In practice, a 30-day script is reduced by half and 15-day supplies are provided twice monthly for a few months until the dosing target is achieved.

#### Dose Rounding

Many drugs used to treat cancer (e.g., chemotherapies) are IV-administered using single-dose vials that are sized in ways that do not always align with commonly administered doses; often, they contain more drug than the patient needs while in other situations many vials are needed to create a full patient dose. Packaging high-cost drugs into single-size vials is very inefficient, both in clinical and financial terms, as the optimal dose typically varies based on a patient’s individual characteristics (e.g., weight, body size). Partially full vials are of little clinical use as they are often in preservative-free formulations that must be used within 6 hours of opening. One-size vial packaging creates $1.5+ billion in avoidable leftover drug waste annually.

*Drug rounding* refers to a technique aimed at lowering the cost of patient care by reducing medication waste and associated costs, without sacrificing efficacy or increasing toxicity. Typically, any calculated dose that falls within 5% to 10% of the established dose is rounded; the rationale being that rounding within this range will not have a negative impact on the safety or effectiveness of the therapy. In practice, dose-rounding options include rounding to the nearest vial size if the rounded dose falls within 10% of the prescribed dose, rounding down to the nearest vial size, and rounding to the nearest vial-size increment (e.g., 50-mg vial for a specific drug). Related methods include syringe increment rounding and rounding to a certain decimal place.

#### Treatment Sequencing / Scheduling

The timing and sequence of a drug’s administration are critical to a treatment’s efficacy and safety. While many traditional cancer therapies are delivered on a strict schedule, the more widespread use of targeted agents employing varied mechanisms of action make it possible to consider different sequencing strategies.

Determining which drugs should be given, in what order or combination, is complicated. Scheduling variables (e.g., concurrent/sequential administration, sequencing order, therapy time, dose intervals, recovery period) can have a consequential effect on a therapy’s efficacy. Due to a paucity of guidelines for optimal sequencing of cytotoxic or targeted agents, treatment sequencing has become a critical issue in oncology research and clinical practice, recognizing the importance of timing for optimizing drug utilization and therapeutic effect.

### Dose Reduction Viability Assessment

The size, packaging, strength, half-life, dose and price of each study drug was reviewed to determine if a reduced dose can be created when common dosing practices are employed. This is largely a factor of variety in a drug’s forms, strengths, dosing frequency, etc. The *variety* of each drug’s possible dosing options was assessed and classified as High (H), Moderate (M) or Low (L), where Moderate denotes *more than adequate* variety. We found that 35 (97%) of the 36 drugs were deemed to offer Medium to High variety, which is more than adequate.

## Appendix B: Pharmaceutical Pricing

Like most products, drugs move through a supply chain to get from the manufacturer to the patient. Unlike other products however, drug prices do not always go up each time they change hands, nor do they often reflect what is actually paid for the drug. As the Congressional Budget Office graphic shows (Figure 5), the pharmaceutical industry involves many buyers and sellers (e.g., maker, wholesaler, distributor, PBM, pharmacy, insurer, physician, patient) and pricing schemes:

- direct retail price (DIRP),
- wholesale acquisition cost (WAC),
- average sale price (ASP),
- average manufacturer price (AMP),
- actual acquisition cost (AAC),
- usual and customary (U&C),
- maximum allowable cost (MAC),
- 340B public health service cost,
- CMS federal upper limit (FUL), and
- *average wholesale price (AWP)*.

**Figure 5:**
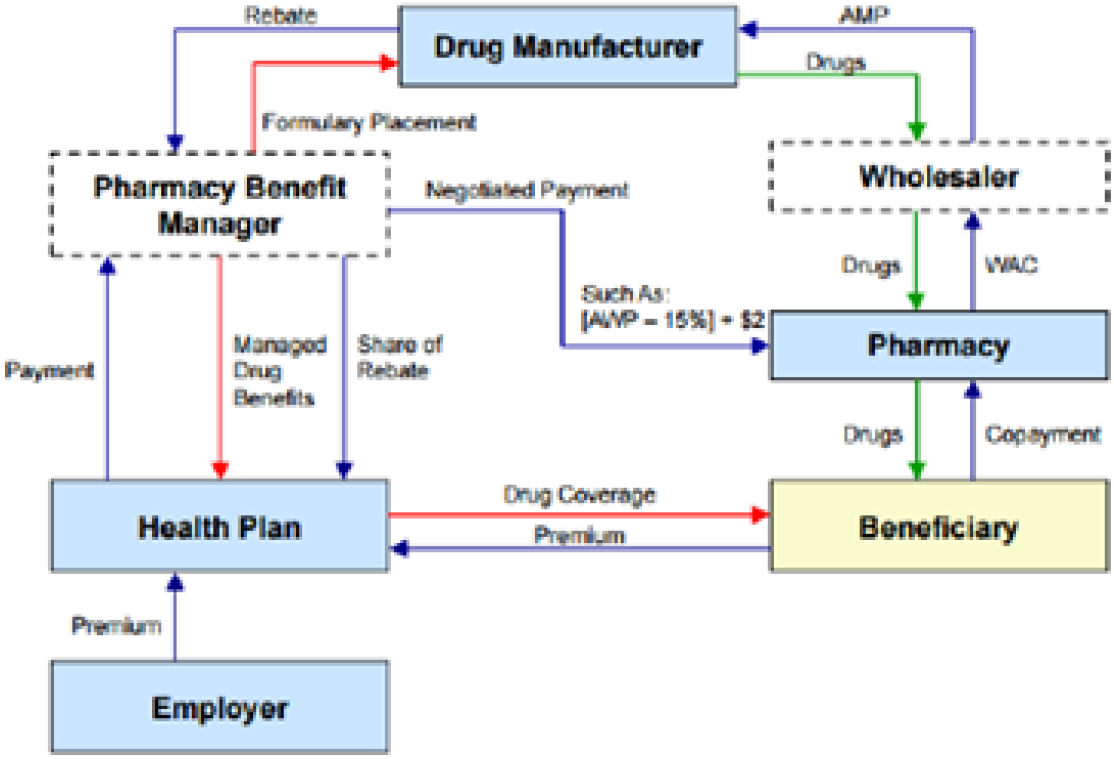
Congressional Budget Office Pharmaceutical Supply Chain

To complicate matters, other factors greatly impact the ultimate cost paid for drugs, including assistance programs (manufacturer coupons, co-pay assistance) and hidden rebates, financial incentives for formulary placement and undisclosed (access, market share, administrative, data) fees that can reach 20% of AWP. The exhibit below shows how drug prices vary based on payment arrangements (pre-fees, rebates, discounts).

AWP, the drug pricing benchmark used by commercial health plans, was used as the price basis for this study; as published online on by Alchemy Health Care Solutions, a CMS-recognized pricing source (on February 2, 2022). It is noteworthy to mention that most manufacturers adjust their drugs’ price, up or down, annually (typically in January or July) as illustrated in Figure 6.

**Figure 6:**
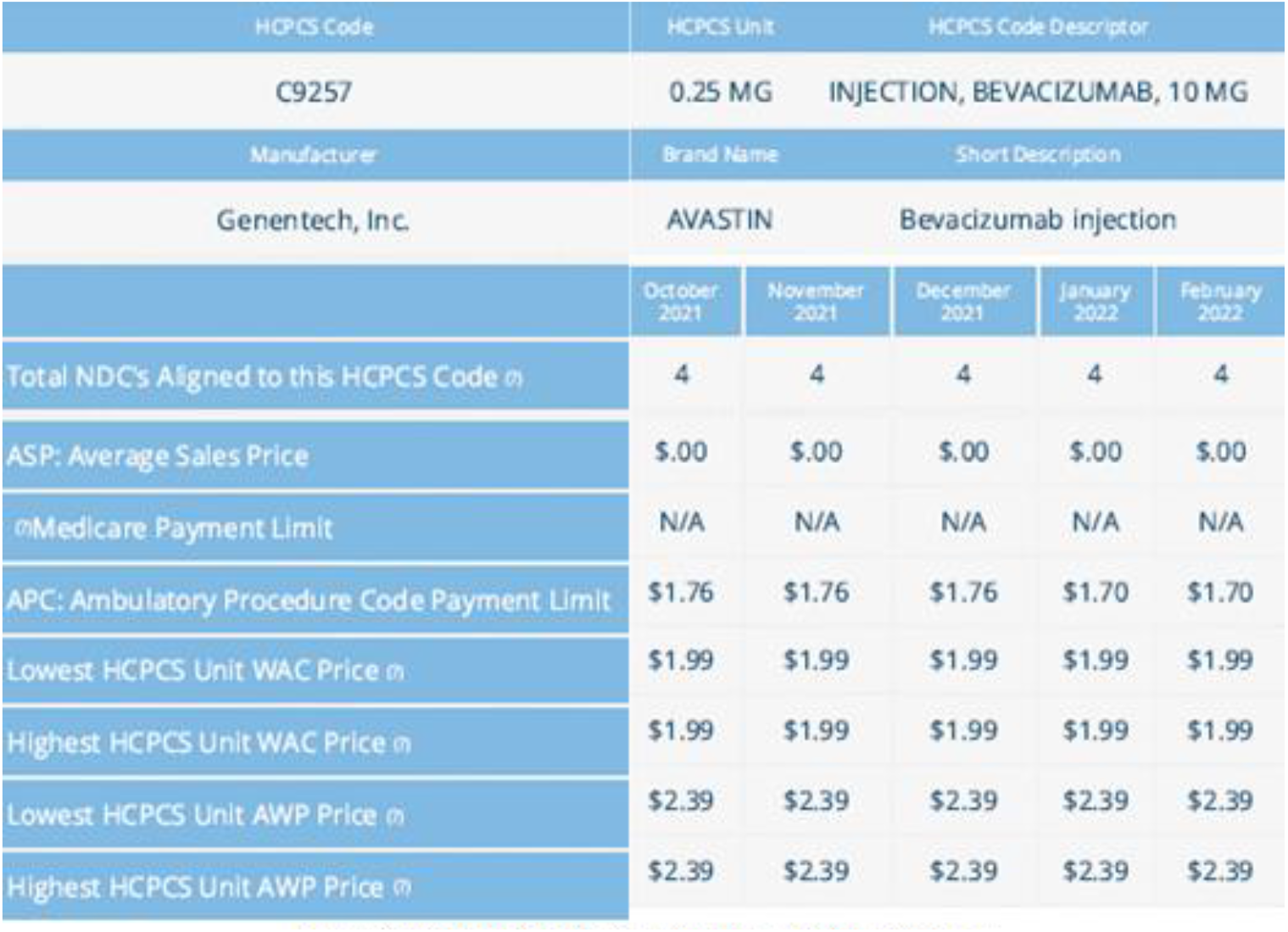
Alchemy Health Care Solutions Price Database

To validate the relative accuracy of the AWP amounts, we cross-checked it against several open-access pharmaceutical pricing databases. Generic drugs were selected, not only based on their cost, but also based on the maker’s reputation.

In those cases where a drug loading dose was recommended, the dose amount used over the 3-month treatment period was used. When patient-specific physical attributes were used to determine dosing, we used the following measures: weight (180 lbs.) height (5’ 10”), body mass index (25 lbs./in^2^), body surface area (1.8 M^2^) and glomerular filtration rate (125 mL/min).

## Notes

### Competing Interest Statement

The Research Consortium serves as a healthcare industry advisor and payer product development consultant to CureMatch, Inc. which provided partial funding for this research.

### Summary of Updates

This version of the manuscript has been updated to increase the "N" from N=5 to N=10 (Case Studies)

